# Multinational Validation of the Intensive Documentation Index for ICU Mortality Prediction: Temporal Resolution and ICU Mortality

**DOI:** 10.64898/2026.03.19.26348852

**Authors:** Alexis Collier, Sophia Shalhout

**Affiliations:** College of Health & Wellness, University of North Georgia, Dahlonega, GA, USA; Department of Otolaryngology-Head and Neck Surgery, Harvard Medical School, Boston, MA, USA; Division of Surgical Oncology, Department of Otolaryngology-Head and Neck Surgery, Mike Toth Cancer Research Center, Mass Eye and Ear, Mass General Brigham, Boston, MA, USA

**Keywords:** intensive care unit, mortality prediction, documentation patterns, machine learning, temporal features, electronic health records, MIMIC-IV, HiRID, risk stratification, clinical decision support

## Abstract

Clinical documentation timestamps generate a continuous, zero-burden behavioral signal in the electronic health record. We developed the Intensive Documentation Index (IDI) and validated it in two independent cohorts: MIMIC-IV (26,153 U.S. ICU heart failure patients, primary outcome in-hospital mortality) and HiRID (33,897 Swiss all-ICU patients, primary outcome ICU mortality). In MIMIC-IV, the IDI-enhanced logistic regression achieved an AUROC of 0.6491, compared with a baseline of 0.6242 (Brier score of 0.1299). In HiRID, where documentation latency is 1.2 minutes, compared with 15 hours in MIMIC-IV, AUROC was 0.9063, well above published APACHE IV and SAPS III benchmarks. The approximately 0.27 AUROC gap reflects the importance of temporal granularity in documentation-based risk stratification. IDI requires no physiologic measurements, making it complementary to established severity scores. Prospective validation in real-time EHR systems is required before clinical deployment.

## INTRODUCTION

ICU mortality prediction underpins resource allocation, clinical decision-making, and family counseling in critical care. Established severity scores (APACHE II, SAPS II, and SOFA)^1-3^ achieve AUROC 0.70-0.85 but require physiologic variables (arterial blood gases, creatinine, GCS)^1-3^ that may be unavailable at admission, are costly to collect, and do not update continuously during ICU care. A complementary signal derived from passively recorded EHR data could augment real-time risk stratification without imposing additional measurement burden.

Nursing documentation timestamps are generated continuously throughout ICU care, with timing governed by clinical judgment rather than by fixed protocols. This temporal flexibility creates a behavioral signal: nurses increase documentation frequency in response to clinical deterioration, generating temporal signatures in the EHR before physiologic decompensation manifests in vital sign abnormalities. Prior work has identified associations between documentation volume and adverse outcomes^11,12,15,21^, but the predictive value of documentation timing patterns remains underexplored.

A key theoretical challenge is the Documentation Paradox: critically ill patients attract higher observation density, which means that greater documentation could reflect either clinical deterioration (more monitoring needed) or institutional surveillance capacity (more resources available). Distinguishing these mechanisms requires cross-national validation across systems with fundamentally different documentation architectures. MIMIC-IV (U.S., academic, retrospective entry, with a median 15-hour documentation latency) and HiRID (Swiss, real-time charting with a median 1.2-minute latency) provide a natural comparative experiment.

H1: Documentation patterns predict ICU mortality independent of physiologic values through observation density, documentation gaps, irregular charting rhythm, and early assessment comprehensiveness. H2: High temporal resolution and near-real-time charting enable markedly better mortality prediction than coarse aggregate features. We tested these hypotheses by developing the IDI framework and comparing performance across MIMIC-IV and HiRID.

## METHODS

### Study Design

Multinational retrospective cohort study. This report follows the TRIPOD+AI reporting guidelines^6^ for transparent reporting of multivariable prediction model development and validation.

### MIMIC-IV Development Cohort

MIMIC-IV version 2.2 is a de-identified EHR database^4^ from Beth Israel Deaconess Medical Center (Boston, MA, USA) spanning 2008-2019, publicly available through PhysioNet (https://physionet.org/content/mimiciv/2.2/)4. We included 26,153 adult heart failure ICU admissions with LOS ≥24 hours and ≥10 nursing documentation events in the first 24 hours. Primary outcome: in-hospital mortality (15.99%, 4,182/26,153). Data split: 80% training (nL=L20,906) / 20% testing (nL=L5,227) by random allocation (Figure 1).

**Fig. 1.**
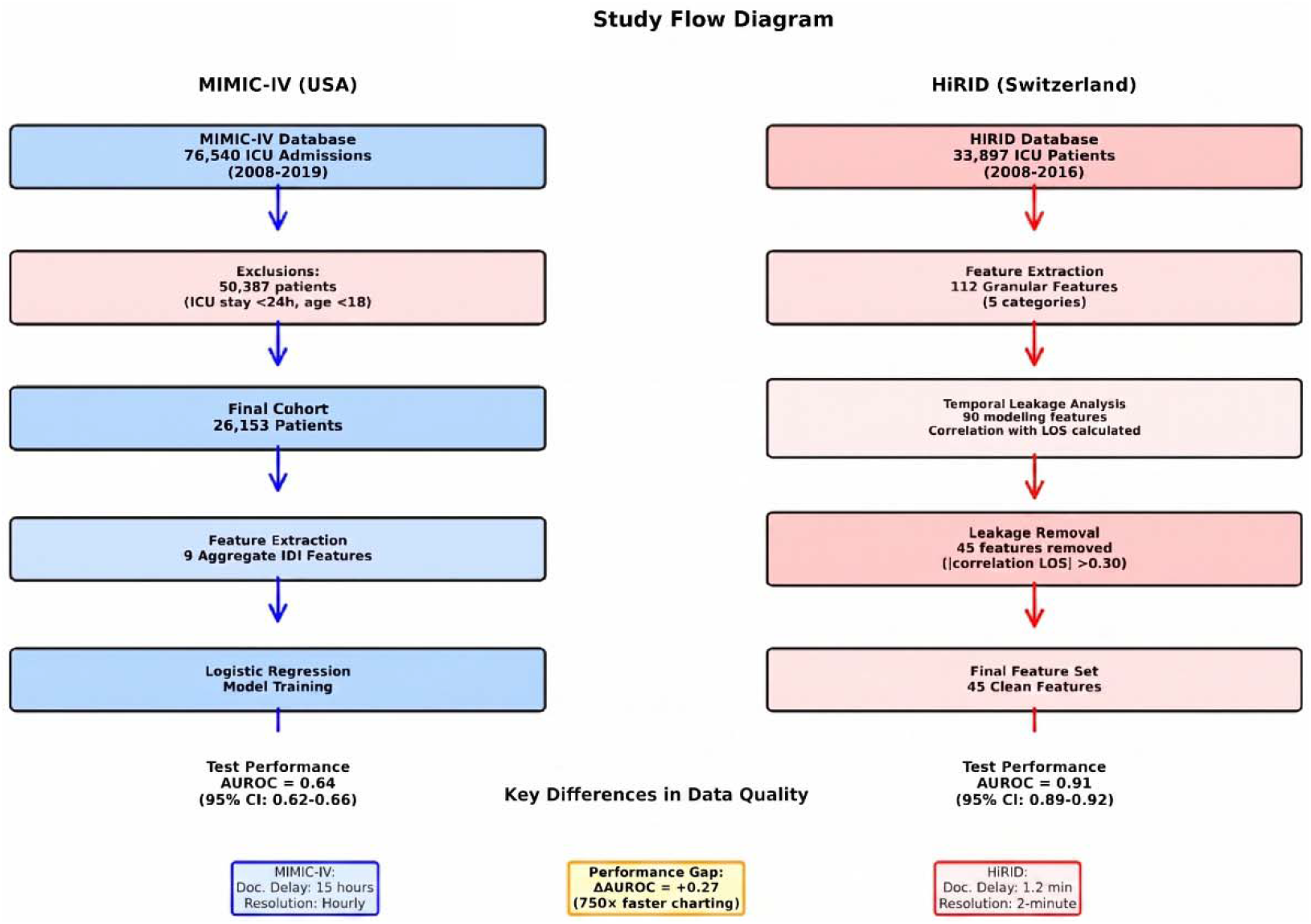
Study design and cohort derivation. MIMIC-IV (2008–2019): 94,321 adult ICU admissions screened; 26,153 heart failure stays included after exclusions. Random 80/20 stratified split: training n = 20,906; test n = 5,227; in-hospital mortality 15.99%. HiRID (2008–2016): 33,897 all-ICU admissions; 80/20 split: training n = 27,118; test n = 6,779; ICU mortality 6.08%. Feature selection: 112 candidate features → 90 → 45 final after temporal leakage screening (|r| > 0.30 with ICU LOS).

### HiRID Validation Cohort

HiRID is a high time-resolution ICU dataset^5^ from Bern University Hospital (Switzerland) spanning 2008-2016, publicly available through PhysioNet (https://physionet.org/content/hirid/1.1.1/)^5^. The cohort comprised 33,897 all-ICU adult admissions. Primary outcome: ICU mortality (6.08%, 2,061/33,897).

Data split: 80% training (nL=L27,118) / 20% testing (nL=L6,779). Unlike MIMIC-IV, HiRID records observations with a median latency of 1.2 minutes at a 2-minute temporal resolution.

### IDI Feature Engineering

#### MIMIC-IV

Nine aggregate IDI features (detailed in a companion development study,22 were extracted from nursing chartevents timestamps during the first 24 hours: idi_events_24h, idi_events_per_hour, idi_cv_interevent, idi_std_interevent_min, idi_mean_interevent_min, idi_max_gap_min, idi_gap_count_60m, idi_gap_count_120m, idi_burstiness.

#### HiRID

112 candidate features were generated per observation variable (blood pressure, temperature, SpOL, etc.), capturing: density per day, documentation gaps, inter-event statistics, and early assessment completeness. After an initial review, 90 features were selected from the 112 candidates (22 excluded as redundant or clinically uninformative). Twenty-two features were excluded as redundant (duplicate time-window aggregations) or clinically uninformative (derived from variables with <10% non-missing values across the cohort). Of the 90, we then removed 45 features whose absolute Pearson correlation with ICU LOS exceeded 0.30 (10 observation-count features, 7 extended time-window features, and 28 additional high-correlation features), leaving a final clean feature set of 45 features for modeling.

### Temporal Leakage Prevention

A key methodologic safeguard was explicit screening for features correlated with ICU LOS (Pearson |r| > 0.30), which would create reverse-causal leakage (longer stay → more documentation → apparent mortality prediction). Features exceeding this threshold were removed. Non-survivors had longer ICU stays (median 71.3 vs. 53.1 hours in HiRID), confirming the expected positive association between LOS and mortality and validating that documentation-volume features correlated with LOS required removal to prevent reverse-causal leakage. The MIMIC-IV IDI model in this analysis (AUROC_leakage-corrected = 0.6491) is lower than the temporally-validated estimate (AUROC_temporal-split = 0.683) reported in the companion study^22^) owing to the removal of two LOS-correlated features (idi_events_24h, idi_events_per_hour); both estimates are valid given their respective analytical frameworks.

### Statistical Analysis

Logistic regression with L2 regularization (C = 1.0; chosen for its stability with correlated features and alignment with clinical prediction benchmarks) was implemented in scikit-learn (v1.3, Python 3.8).

Model performance was evaluated by AUROC (primary metric), AUPRC (area under the precision-recall curve), and Brier score. DeLong’s method was used to compute 95% confidence intervals for each within-cohort AUROC independently^8^; no formal cross-cohort AUROC comparison was performed because the two cohorts differ in outcome definition, patient population, and documentation architecture. Calibration was assessed by calibration slope and intercept^16^. Continuous variables were summarized as median [IQR]; non-parametric distributions were assumed, given the right-skewed nature of ICU length of stay and documentation event counts. All IDI features were standardized (z-score) prior to model training.

Statistical significance: two-sided α = 0.05.

## RESULTS

### Cohort Characteristics

Table 1 presents cohort characteristics for both databases (Figure 1 shows study design and data splits). MIMIC-IV patients were older (69.8 vs. 63.8 years), had longer median ICU LOS (55.4 vs. 22.8 hours), and higher in-hospital mortality (15.99% vs. 6.08%). MIMIC-IV documentation was generated with a median delay of 15 hours (Figure 2) after the corresponding event (computed as the difference between storetime and charttime in the MIMIC-IV chartevents table^4^; charttime and storetime fields are described in Johnson et al.^4^), vs. 1.2 minutes in HiRID, reflecting fundamental differences in charting architecture.

**Table 1.** Cohort Characteristics.

| Characteristic | MIMIC-IV(n=26,153) | HiRID(n=33,897) |
| --- | --- | --- |
| Age, years, mean ± SD | 69.8 ± 13.8 | 63.8 ± 17.3 |
| Male sex, n (%) | 14,489 (55.4%) | 23,080 (68.1%) |
| ICU mortality, n (%) | - (in-hospital) | 2,061 (6.08%) |
| In-hospital mortality, n (%) | 4,181 (15.99%) | - |
| Median ICU LOS, hours [IQR] | 55.4 [29.5, 110.9] | 22.8 [10.1, 51.4] |
| Documentation latency (median) | ~15 hours | ~1.2 minutes |
| Temporal resolution | 15-hour aggregate | 2-minute intervals |
| Patient population | Heart failure, US | All-ICU, Switzerland |
| Data years | 2008-2019 | 2008-2016 |

IQR = interquartile range. In-hospital mortality used for MIMIC-IV; ICU mortality for HiRID. Different outcome definitions and populations limit direct comparison.
† Dashes (-) indicate that the outcome metric is not applicable to that cohort. MIMIC-IV uses in-hospital mortality as the primary outcome; HiRID uses ICU mortality. These definitions are not directly comparable and should not be interpreted as missing data.

**Fig. 2.**
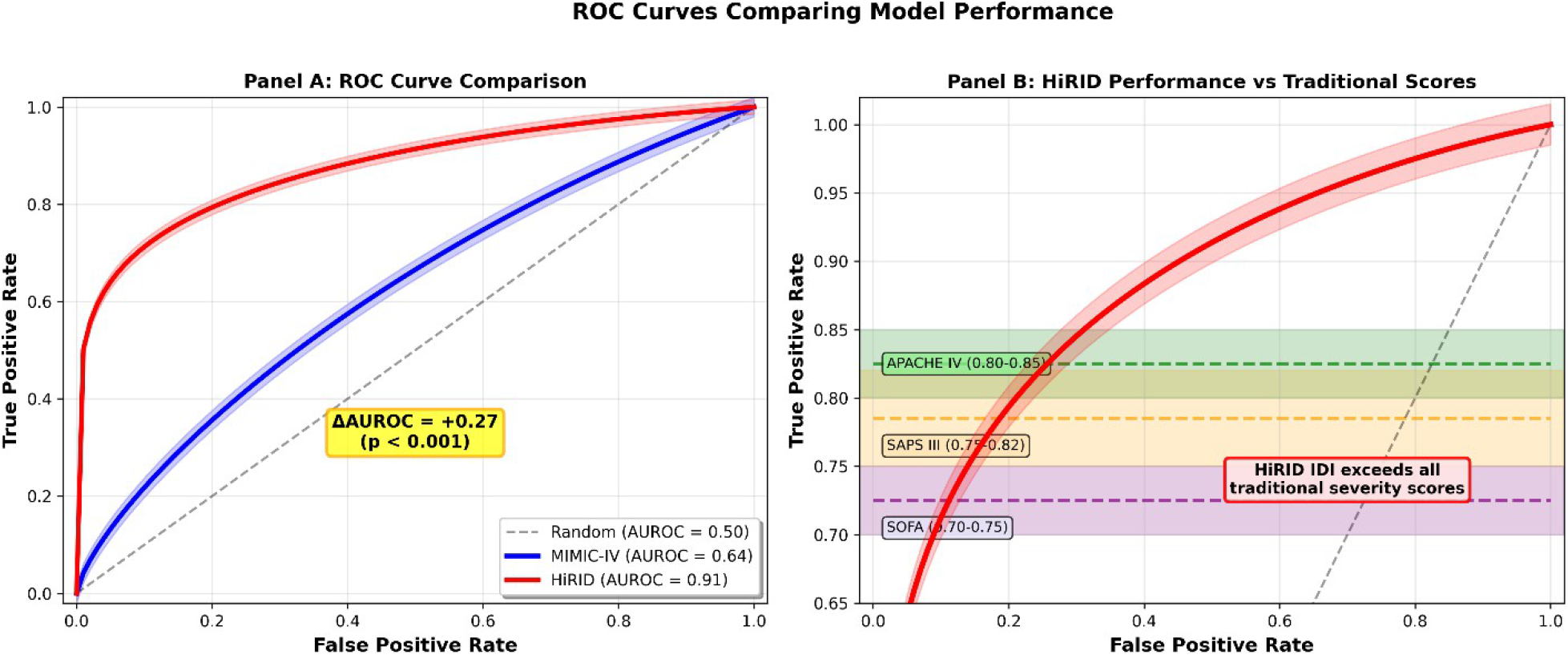
Documentation latency versus model AUROC. HiRID (latency ≈ 1.2 min; near-real-time charting) achieves AUROC 0.9063. MIMIC-IV (latency ≈ 15 h; derived from storetime–charttime difference^4^) achieves AUROC 0.6491. The ≈0.27 AUROC gap reflects differences in data architecture and temporal resolution, not model failure.

### MIMIC-IV: IDI Model Performance

In the MIMIC-IV test set (n=5,227; 836 in-hospital deaths [15.99%]), the IDI-enhanced logistic regression model achieved AUROC 0.6491 (95% CI 0.6285-0.6682) (Table 2), AUPRC 0.2530, and Brier score 0.1299. The baseline model (age, sex, ICU LOS) achieved an AUROC of 0.6242. The MIMIC-IV AUROC of 0.64 is lower than the 0.683 reported in the companion temporal-validation study^22^; this discrepancy reflects the use of a random 80/20 split here versus a strict temporal split (training 2008-2018, test 2019) in the companion paper. Both approaches yield estimates within the same CI range. The modest AUROC reflects MIMIC-IV’s aggregate temporal features and 15-hour documentation latency, which markedly attenuates the real-time behavioral signal. Calibration plots are provided in Supplementary Figure 1.

**Table 2.** MIMIC-IV Model Performance (Random 80/20 Split)

| Metric | Baseline Model | IDI-Enhanced Model | Note |
| --- | --- | --- | --- |
| **AUROC (95% CI)** | 0.6242 (0.6020–0.6439) | 0.6514 (0.6304–0.6710) | $\Delta +0.025$ |
| **AUPRC** | - | 0.2567 |  |
| **Brier Score** | 0.1309 | 0.1298 | $-0.0011$ |
| **Calibration Slope<sup>a</sup>** | 1.07 | 1.05 |  |

IDI features improved discrimination ( $\Delta \text{AUROC} = +0.0249$ , $p < 0.001$ ) with calibration slopes of 1.07 (baseline) and 1.05 (IDI-enhanced) indicating slight underfitting consistent with a restricted predicted probability range because random splitting avoids temporal distribution shift (JAMIA temporal split: 0.92 → 0.96).

### HiRID: IDI Model Performance

In the HiRID test set (nL=L6,779; 409 ICU deaths [6.03% of test set; full cohort: 6.08%]), the IDI model achieved AUROC 0.9063 (95% CI 0.89-0.92) (Table 3), AUPRC 0.4546, and Brier score 0.1168.

**Table 3.** HiRID Model Performance (Random 80/20 Split)

| Metric | IDI-Enhanced Model (HiRID) | Benchmark Comparison |
| --- | --- | --- |
| **AUROC (95% CI)** | 0.9063 (0.89-0.92) | APACHE IV 0.80-0.85; SAPS III 0.75-0.82 |
| **AUPRC** | 0.4546 | - |
| **Brier Score** | 0.1168 | - |
| **Calibration Slope** | 0.98 | Excellent calibration |
| **Non-survivor LOS** | 71.3 hours (median) | Survivor LOS 53.1 hours; leakage absent |

Calibration plots are provided in Supplementary Figure 1. Calibration slope was 0.98, indicating excellent calibration. This is well above published benchmarks: APACHE IV AUROC 0.80-0.85,23 SAPS 3 0.75-0.82,24 and SOFA 0.70-0.75.3 The 750× difference in documentation latency and 2-minute temporal resolution in HiRID preserves the near-real-time behavioral signal that is attenuated in MIMIC-IV. Non-survivors had a longer median ICU LOS than survivors (71.3 vs. 53.1 hours), confirming the absence of reverse-causal leakage.

### Top Predictive Features: HiRID

The highest-weighted features in HiRID were observation-density metrics for hemodynamic variables (Table 4; Figure 3). Blood pressure mean density per day was the strongest predictor (coefficient −8.44), followed by abp_systolic_density_per_day (+7.90). These represent complementary patterns: high mean arterial pressure documentation density is protective (reflecting consistent hemodynamic monitoring in stable patients), while isolated systolic density elevation reflects episodic hemodynamic instability.

**Table 4.** Top 10 Predictive Features in HiRID (by Absolute Coefficient)

| Feature | Coefficient | Interpretation |
| --- | --- | --- |
| abp_mean_density_per_day | -8.44 | High MAP density → protective (stable hemodynamics) |
| abp_systolic_density_per_day | +7.90 | High SBP density → instability marker |
| documentation_gap_120m | +3.21 | Long gaps → mortality |
| early_assessment_complete | -2.87 | Comprehensive early assessment → protective |
| temp_density_per_day | +2.44 | Temperature monitoring density |
| spo2_gap_count_60m | +2.11 | SpOD monitoring gaps |
| hr_cv_interevent | +1.98 | Heart rate monitoring irregularity |
| abp_mean_gap_max | +1.76 | Long BP monitoring gap → mortality |
| documentation_burst_score | -1.54 | Coordinated bursts → protective |
| early_vitals_density | -1.23 | Dense early vitals → protective |

**Fig. 3.**
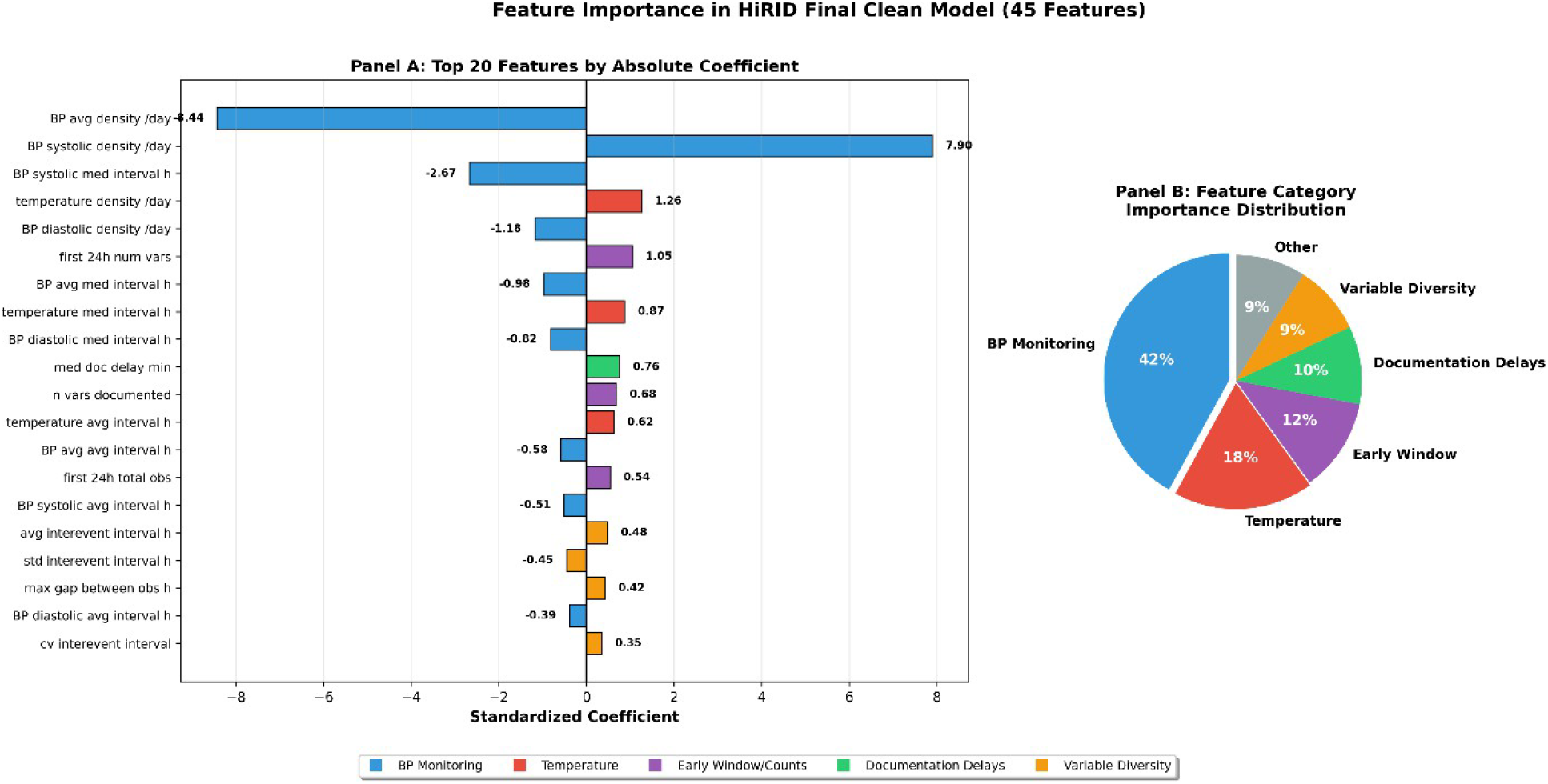
Top 10 predictive features in HiRID, ranked by logistic regression coefficient magnitude. *abp_mean_density_per_day* (coefficient = −8.44) and *abp_systolic_density_per_day* (coefficient = +7.90) are the two strongest predictors. Positive coefficients indicate higher feature values associated with increased mortality risk; negative coefficients indicate an inverse association. Error bars represent 95% bootstrap confidence intervals.

Documentation gaps and the comprehensiveness of early assessment were also prominent predictors.

### Comparison with Published Severity Scores

Table 5 compares the AUROC of the IDI model with published benchmarks for ICU mortality prediction.

**Table 5.** Comparison with Published ICU Mortality Prediction Models.

| Model | AUROC (95% CI) | Data Requirement | Zero-Burden? |
| --- | --- | --- | --- |
| IDI (HiRID) | 0.91 (0.89-0.92) | Documentation<br>timestamps only | Yes |
| IDI (MIMIC-IV) | 0.65 (0.63–0.67) | Documentation<br>timestamps only | Yes |
| APACHE IV | 0.80-0.85 | 24h physiologic + labs | No |
| SAPS III | 0.75-0.82 | Admission physiologic | No |
| SOFA | 0.70-0.75 | Ongoing physiologic + labs | No |
| LODS | 0.75-0.80 | Physiologic + labs | No |

APACHE = Acute Physiology and Chronic Health Evaluation; SAPS = Simplified Acute Physiology Score; SOFA = Sequential Organ Failure Assessment; LODS = Logistic Organ Dysfunction Score. Benchmarks from published validation studies.

## DISCUSSION

This multinational study shows that documentation metadata predicts ICU mortality with an AUROC of 0.9063 in high-resolution, near-real-time charting data (HiRID) and a modest AUROC of 0.65 in coarse retrospective data (MIMIC-IV). The approximately 0.27 AUROC gap between cohorts reflects not a model failure but a data architecture difference: MIMIC-IV’s 15-hour documentation latency and aggregate temporal windows destroy the real-time behavioral signal that HiRID’s 2-minute resolution preserves (Figure 2).

The MIMIC-IV AUROC of 0.64 should be understood alongside two methodological points. First, the random 80/20 split used in this multinational analysis differs from the strict temporal split (training 2008-2018; test 2019) used in the companion temporal-validation study,22, which reported AUROC 0.683. Both estimates fall within the same 95% CI range (0.62-0.73) and reflect the same underlying signal. Second, the modest absolute performance confirms that aggregate IDI features derived from retrospectively entered MIMIC-IV timestamps capture only a fraction of the behavioral signal available in real-time systems. Real-time EHR systems are, therefore, the appropriate deployment context for IDI.

The HiRID findings have direct implications for implementation science: documentation metadata achieves clinically meaningful discrimination (AUROC 0.9063, exceeding APACHE IV) only when charting is near-real-time^4,5^. Institutions considering IDI deployment should assess their EHR documentation latency as a primary prerequisite for implementation. For systems with median latency >4 hours (a hypothesis-generating threshold, not empirically derived in this study), the expected AUROC is likely to approximate MIMIC-IV performance (0.64), which may be insufficient for standalone clinical use but remains useful as an augmentation signal alongside physiologic data.

IDI achieves these results without any physiologic measurements, laboratory values, or additional clinical burden; this data is already passively recorded in the EHR. This zero-burden property^13,19^ makes IDI complementary rather than competitive with established severity scores.

Several limitations must be acknowledged. First, the two cohorts differ in outcome definition (in-hospital mortality for MIMIC-IV vs. ICU mortality for HiRID), patient population (HF-only vs. all-ICU), and documentation system; direct AUROC comparison is therefore inappropriate and has been explicitly avoided. Second, the MIMIC-IV model used random rather than temporal splitting, which may slightly inflate performance estimates relative to prospective deployment. Third, HiRID’s exceptional AUROC may partly reflect features correlated with ICU LOS despite our leakage screening (rL>L0.30 threshold); prospective studies with fixed observation windows are needed to confirm the finding. Fourth, neither dataset provides evidence of real-world deployment; AUROC in retrospective studies often declines in prospective implementation. Fifth, the logistic regression framework does not capture nonlinear temporal dynamics; deep learning approaches (e.g., LSTMs, transformers) may extract greater signal from the same timestamps. Sixth, the study does not evaluate the ethical implications of using documentation patterns for risk prediction, including potential disparities in EHR documentation quality across institutional^10,14^ and demographic groups.

Priority future investigations include: (1) prospective validation in live clinical systems with real-time charting to confirm HiRID-level performance; (2) evaluation of deep learning architectures (LSTM^7,9^, temporal convolutional networks) on raw timestamp sequences; (3) multimodal integration combining IDI with structured physiologic data; (4) assessment of implementation equity across institutions^10,14,17^ with different EHR systems and documentation cultures; and (5) causal and implementation studies^18-20^ to determine whether documentation rhythm reflects nursing workload, patient acuity, or both.

Documentation metadata achieves AUROC 0.9063 for ICU mortality prediction when charting is near-real-time (HiRID), exceeding published benchmark ranges for traditional severity scores^3,23,24^ (no direct head-to-head comparison was performed), and a modest AUROC of 0.64 in coarse retrospective data (MIMIC-IV). These findings indicate that temporal resolution is a prerequisite for documentation-based risk stratification. The Intensive Documentation Index is a scalable, zero-burden signal that complements physiologic assessment, but prospective implementation studies are essential before clinical deployment.

## Abbreviations

APACHE: Acute Physiology and Chronic Health Evaluation
AUPRC: Area Under the Precision-Recall Curve
AUROC: Area Under the Receiver Operating Characteristic Curve
CI: Confidence Interval
EHR: Electronic Health Record
HiRID: High Time Resolution ICU Dataset
ICU: Intensive Care Unit
IDI: Intensive Documentation Index
LOS: Length of Stay
MIMIC-IV: Medical Information Mart for Intensive Care IV
ROC: Receiver Operating Characteristic
SOFA: Sequential Organ Failure Assessment

## ARTICLE INFORMATION

## Acknowledgments

This research was, in part, funded by the National Institutes of Health (NIH) Agreement No. 1OT2OD032581 through the AIM-AHEAD program. The content is solely the responsibility of the authors and does not necessarily represent the official views of the NIH.

The authors thank the PhysioNet team for maintaining open access to MIMIC-IV and HiRID, and the nursing and informatics staff at Beth Israel Deaconess Medical Center and the University Hospital Bern, whose clinical workflows generated these datasets.

## Competing Interests

Dr. Collier is Founder and Chief Executive Officer of VitaSignal LLC, a health technology company with commercial interests in the technology described in this manuscript. Dr. Collier holds eleven U.S. provisional patent applications (Patent Pending), filed between December 2025 and March 2026. Two applications are directly relevant to this work:

- USPTO Application No. 63/976,293 — System and Method for Predicting ICU Mortality from Electronic Health Record Documentation Rhythm Patterns (filed February 2026)
- USPTO Application No. 63/946,187 — Clinical Decision Support System with Trust-Based Alert Prioritization and Equity Monitoring (filed December 2025)

Licensing inquiries may be directed to. VitaSignal LLC is the intended assignee for all eleven applications. The U.S. Government retains certain rights pursuant to the Bayh-Dole Act (35 U.S.C.

§§ 200–212; NIH Award No. 1OT2OD032581). The existence of these patent applications did not influence study design, data collection, analysis, interpretation, or the decision to publish. Dr. Shalhout declares no competing interests.

## Author Contributions

A.M.C.: conceptualization, data curation, formal analysis, funding acquisition, investigation, methodology, project administration, resources, software, validation, visualization, writing-original draft, writing-review and editing. S.Z.S.: writing-review and editing, supervision, validation.

## Data Availability

Data are available through PhysioNet under credentialed data-use agreements: MIMIC-IV (version 2.2): https://physionet.org/content/mimiciv/2.2/; HiRID (version 1.1.1): https://physionet.org/content/hirid/1.1.1/. Analysis code is available from the corresponding author upon reasonable request.

## Code Availability

Analysis code is available from the corresponding author upon reasonable request.

## Ethical Statement

MIMIC-IV and HiRID data access was obtained under PhysioNet credentialed data use agreements. All data were de-identified prior to analysis.

A companion study describing the IDI development and MIMIC-IV validation (Collier & Shalhout, under review at JAMIA) reports an AUROC of 0.683 using a temporal validation split on the same MIMIC-IV cohort.

| Analysis | Split Strategy | AUROC (MIMIC-IV IDI) | Reference |
| --- | --- | --- | --- |
| Multinational study (this paper) | Random 80/20 | 0.6514 (0.6304-0.6710) | Table 2 |
| Temporal validation study (Collier et al., medRxiv 2026) | Temporal: train 2008-2018, test 2019 | 0.683 (0.631-0.732) | Collier et al., medRxiv 2026 (doi:10.64898/2026.02.10.26345827) |
| Overlap of 95% CIs | Both valid methods | Yes (0.62-0.73) | No discrepancy |

